# Global transmission network of SARS-CoV-2: from outbreak to pandemic

**DOI:** 10.1101/2020.03.22.20041145

**Authors:** Pavel Skums, Alexander Kirpich, Pelin Icer Baykal, Alex Zelikovsky, Gerardo Chowell

## Abstract

**Background:** The COVID-19 pandemic caused by the severe acute respiratory syndrome coronavirus 2 (SARS-CoV-2) is straining health systems around the world. Although the Chinese government implemented a number of severe restrictions on people’s movement in an attempt to contain its local and international spread, the virus had already reached many areas of the world in part due to its potent transmissibility and the fact that a substantial fraction of infected individuals develop little or no symptoms at all. Following its emergence, the virus started to generate sustained transmission in neighboring countries in Asia, Western Europe, Australia, Canada and the United States, and finally in South America and Africa. As the virus continues its global spread, a clear and evidence-based understanding of properties and dynamics of the global transmission network of SARS-CoV-2 is essential to design and put in place efficient and globally coordinated interventions.

**Methods:** We employ molecular surveillance data of SARS-CoV-2 epidemics for inference and comprehensive analysis of its global transmission network before the pandemic declaration. Our goal was to characterize the spatial-temporal transmission pathways that led to the establishment of the pandemic. We exploited a network-based approach specifically tailored to emerging outbreak settings. Specifically, it traces the accumulation of mutations in viral genomic variants via mutation trees, which are then used to infer transmission networks, revealing an up-to-date picture of the spread of SARS-CoV-2 between and within countries and geographic regions.

**Results and Conclusions:** The analysis suggest multiple introductions of SARS-CoV-2 into the majority of world regions by means of heterogeneous transmission pathways. The transmission network is scale-free, with a few genomic variants responsible for the majority of possible transmissions. The network structure is in line with the available temporal information represented by sample collection times and suggest the expected sampling time difference of few days between potential transmission pairs. The inferred network structural properties, transmission clusters and pathways and virus introduction routes emphasize the extent of the global epidemiological linkage and demonstrate the importance of internationally coordinated public health measures.

## 1 Introduction

The COVID-19 pandemic due to the SARS-CoV-2 virus that emerged out of the city of Wuhan, Hubei Province in China in December, 2019 [15, 31, 71, 46, 41, 66, 53, 16], is now straining or overwhelming health care systems around the world [1]. As of March 2020, hundreds of new confirmed cases have been reported daily in multiple countries of every continent [54, 51, 59, 35, 30] [13, 17, 39] while the global death toll has passed 13,000. To combat the spread of the virus, in the absence of a vaccine or specific treatments, an increasing number of nations are putting in place social distancing interventions ranging from school closures, quarantine orders on segments of the population, and banning large public gatherings. In order to devise effective control strategies at different spatial scales, it is critical to have a clear understanding of transmission pathways of SARS-CoV-2, including local human-to-human transmission dynamics [31, 12, 49, 46, 37] as well as long-range transmission events (country-to-country) [65]. To characterize the origin, geographic extent and epidemiological parameters of the epidemic, phylogenetics and phylodynamics inference tools have proved useful during this and past epidemic emergencies [68, 69, 38, 61, 10, 63, 11, 10, 5, 48, 70, 42, 60, 67, 19]. However, previous studies indicate that methods based on genetic networks are more powerful and efficient to identify transmission patterns and ascertain transmission links compared to methods based on binary phylogenies [62].

In this study, we used a network-based approach to analyze the global SARS-CoV-2 transmission patterns by constructing the transmission network based on genomic compositions of viral sequences sampled around the world and before the pandemic declaration on March 11, 2020. Here we sought to characterize the transmission pathways that facilitated the virus to establish itself as a pandemic. Such analyses rely on viral genomic data that continues to accumulate in near real time as next-generation sequencing technologies are now more widely used. The richness and accessibility of the genomic data distinguishes this SARS-CoV-2 pandemic from previous large-scale epidemics or pandemics including the 2009/AH1N1 influenza pandemic and the 2003 SARS outbreaks. This provides an unprecedented opportunity to gain insights into the epidemiological and evolutionary dynamics of this emerging virus spreading in an essentially naive population. Another feature of the scope of the genomic data for COVID-19 is the high sampling density available to investigate the early transmission stages. Indeed, although the virus genetic diversity gradually increases as the virus spreads, particular genomic variants have been repeatedly sequenced at different time points and geographical locations. This indicates that the available sequencing data cover a significant part of the evolutionary space explored from the onset of the epidemics. In this setting, viral evolution could be accurately modeled, reconstructed and visualized by genetic networks models [8, 52], which have been successfully used for the analysis of different viral epidemics [52, 62, 9]. However, in emerging outbreak settings, when all circulating viral genomes are relatively close to each other, methods that allow the analysis of viral population structures at finer resolution than provided by state-of-the-art network models are needed. We achieve such resolution by using mutation trees [34] (related to character-based phylogenies [24]) that keep track of the accumulation of mutations in viral populations. Moreover, as the SARS-CoV-2 population contain relatively few 4-gamete rule violations, it is feasible to efficiently generate and analyze all plausible mutation trees. These trees, in turn, allow for accurate inferences of transmission networks.

Our results presented here summarize the up-to-date picture of the spread of SARS-CoV-2 between and within countries and geographic regions. The structural properties of the inferred network, transmission clusters and pathways as well as virus introduction routes emphasize the extent of the global epidemiological transmission network. It also demonstrates the importance of internationally coordinated public health measures and highlights how epidemiological and molecular surveillance analyses complement each other to characterize the spatial-temporal spread of epidemics.

## 2 Methods

### 2.1 Data preprocessing

We obtained the genomics data and associated metadata for this study from the Global Initiative on Sharing All Influenza Data (GISAID) database [56] that hosts genetic datasets for SARS-CoV-2, which have been self-reported from multiple sources. The sequences identified by GISAID as low-quality have been removed from consideration. The reference genome was identified and taken from the literature [64]. It coincides with the most prevalent sequence from GISAID sampled from Wuhan and a number of other locations. The remaining sequences were aligned to the consensus using MUSCLE [20] and trimmed to the same length, yielding n=319 aligned sequences of length 29772 base pairs (bp). In order to be as conservative as possible in the mutation calling, gaps and non-identifiable positions have been assumed to have major alleles. Next, genomic positions without variation have been removed, leaving *m* = 274 single nucleotide variants (SNVs) for further analysis. Finally, the alignment was represented by the *n × m* (0, 1)-mutation matrix *M* with rows corresponding to sequences and columns corresponding to SNVs, where *M*_*i,j*_ = 1 whenever the *i*-th genome has a minor allele at the position *j* with respect to the reference.

### 2.2 Dimensionality reduction and clustering

Viral sequences were embedded into the 2-dimensional space using T-distributed Stochastic Neighbor Embedding (t-SNE) [43] based on pairwise hamming distances between aligned sequences. The embedding points were clustered by k-means clustering, and the optimal number of clusters was estimated using the gap statistics [58].

### 2.3 Mutation tree reconstruction

In a mutation tree,

- internal nodes correspond to mutations (columns of the mutation matrix), with the root representing the zero mutation (the absence of mutations);
- leafs represent sampled genomes (rows of the mutation matrix)
- mutational profile of each sequence consists of mutations on the path from the corresponding leaf to the root.

Note that this tree does not have to be binary. In the perfect phylogeny model, which is the most widely used character-based phylogenetic model [25], each mutation can be represented by a single internal node implying that each mutation occurs only once. The perfect phylogeny model can only explain the data without 4-gamete rule violation, i.e. whenever for each pair of columns in the mutation matrix *M*, there are no 4 sequences that have all possible combinations of alleles (0, 0), (0, 1), (1, 0), (1, 1) at that positions. The sequencing data accumulated for SARS-CoV-2 includes several 4-gamete rule violations, thus implying repeated mutations in the same genomic positions. Therefore, instead of using the perfect phylogeny model, we fit the data to Camin-Sokal phylogenetic model, according to which repeated mutations are allowed, mutation losses are not allowed and each mutation could be acquired at most twice [7].

We first identify potential repeated mutations and then construct plausible mutations trees taking into account for the possibility of false SNV calls resulting from the sequencing noise as follows:

#### 1) Identify potential repeated mutations

This is achieved in a most parsimonious way using a graph *G*_4*g*_, whose vertices are SNVs, and two vertices are adjacent whenever the corresponding pair of SNVs violates 4-gamete rule. If we remove the SNVs corresponding to vertices of *G*_4*g*_, then the remaining mutation matrix *M* can be explained without any repeated mutations. Therefore, we are looking for the the minimum vertex cover of *G*_4*g*_, i.e. the minimum number of vertices in *G*_4*g*_ whose removal destroys all edges in *G*_4*g*_. The set of genomic positions corresponding to vertices in the minimum vertex cover of *G*_4*g*_ forms the most parsimonious set of mutations that should be repeated. We find all minimal vertex covers using Bron-Kerbosch algorithm [6] for maximum independent sets generation; this approach uses the fact that complements of maximum independent sets are exactly minimum vertex covers. Since the number of 4-gamete rule violations for SARS-CoV-2 data is relatively small, this method is fast and allows to significantly simplify the phylogeny reconstruction.

#### 2) Construct mutation trees

For each minimum set *R* = {*m*_1_, …, *m*_*k*_} of potential repeated mutations found in the previous step, we construct a character-based Camin-Sokal phylogeny which minimizes the number of mismatches with the original mutation matrix *M* as follows. We generate an extended set of mutations *P* = {1, …, *m, m* + 1, …, *m* + *k*}, where each original mutation *j ∈* {1, …, *m*} *¥ R* is represented by a single copy and each mutation *j ∈ R* is represented by a pair of copies *C*(*j*). The sought-for Camin-Sokal phylogeny *T* would be a perfect phylogeny with respect to the extended set of mutations *P*. To construct this phylogeny and the corresponding extended mutation matrix *X*, we utilize an integer linear programming (ILP) approach of Dan Gusfield [26]. The following binary variables are used:

a. *X*_*i,j*_ = 1 whenever the genomic variant *i* has a mutation *j* from the extended set *P, i* = 1, ‥, *n*; *j* = 1, …, *m* + *k*.
b. *D*_*j,l,a,b*_ = 1 whenever there is a sequence that have an allele combination (*a, b*) at the positions *j* and *l, j* = 1, …, *m* + *k*; *l* = *j* + 1, …, *m* + *k*; (*a, b*) *∈* {(0, 0), (0, 1), (1, 0), (1, 1)}. Then we seek to minimize the total number of mismatches between the observed mutation matrix M and the mutation profiles defined by the tree T by minimizing the objective function

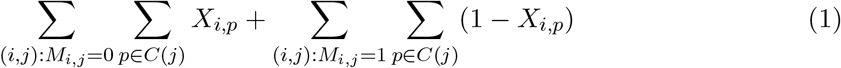

subject to constraints

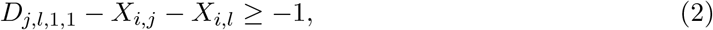

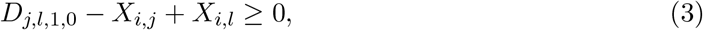

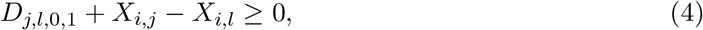

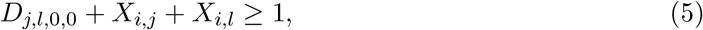

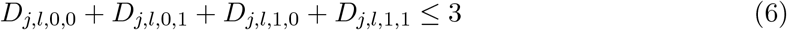

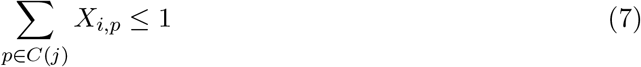

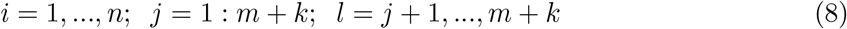

The first 4 sets of constraints enforces the relations between the variables *X*_*i,j*_ and *D*_*i,j,a,b*_ specified by (a) and (b), the fifth set of constraints guarantees that *T* is the perfect phylogeny with respect to the extended set of mutations *P*, and the last set of constraints ensures that two gains of the same mutation appear only in parallel lineages. The instances of the ILP problem were solved to optimality using Gurobi 8.1. (Gurobi Optimization, LLC [47]). The trees were constructed for all potential sets of repeated mutations, and the tree with the best objective function was selected.

### 2.4 Transmission network construction and bootstrapping

The transmission network defined by the mutation tree *T* is a directed graph, whose vertices represent viral genomes, and two genomes are connected by an arc if their mutational composition suggests potential direct or indirect transmission linkage between their hosts. For a given mutation tree *T*, the corresponding transmission network *G*_*T*_ is constructed as follows:

1. Collapse sequences that share the same parent in *T* into a single *haplotype*. The set of haplotypes forms the vertex set of *G*_*T*_.
2. A pair of haplotypes *h*_*i*_ and *h*_*j*_ are connected by a directed arc whenever (a) the parent of *h*_*i*_ is an ancestor of the parent of *h*_*j*_ and (b) there is no haplotype *h*_*k*_ whose parent belongs to the path between the parents of *h*_*i*_ and *h*_*j*_.

In graph-theoretical terms, *G*_*T*_ is the transitive reduction of the reachability graph of the parents of observed haplotypes.

To quantify the uncertainty for the hypothesized transmission links, bootstrapping of the transmission networks was performed. At each bootstrap, *m* mutations were sampled with replacement from the the original set of mutations, and the mutation tree and the transmission network were constructed using the obtained mutation matrix as discussed above. For each potential transmission link *e*, its bootstrap probability *p*_*e*_ was calculated. The final consensus transmission tree was estimated as the maximum-weight spanning arborescence [18, 21] with respect to the weights *p*_*e*_.

## 3 Results

### 3.1 Transmission clusters

t-SNE plots combined with *k*-means clustering suggest that as of March 11, 2020 five distinct viral subpopulations have been circulating globally (Fig. 1). These subpopulations could be roughly classified as follows:

1. The original cluster(shown in black) that includes sequences sampled from Wuhan and other mainland China provinces during the early transmission phase, as well as from USA, Singapore, Taiwan, Thailand, South Korea, Nepal and several European countries are most probably epidemiologically linked with China at earlier outbreak stages.
2. European cluster (shown in red) that includes sequences almost exclusively from European countries, as well as from a few non-European countries (Nigeria, Mexico, Brazil) most of whom are documented to be epidemiologically linked to Europe.
3. The cluster from mostly Pacific countries (shown in blue) that includes sequences from mainland China, South Korea, Hong Kong, Singapore, Vietnam, Australia, USA and Chile, including the subcluster of sequences from the US state of Washington corresponding to the ongoing outbreak there.
4. The cluster that include significant portion of genomes sampled in Australia and New Zealand (green). It should be noted that 2 infected hosts from Australia and 1 infected host from New Zealand are reported to have travel history to Iran. It may mean that this strain is a branch of an Iranian strain tied to the epidemics at a much larger scale in that country, and two sequences from China that belong to this cluster are related to their common ancestor. However, full-length sequences from Iran are currently unavailable, and therefore this scenario is currently hypothetical.
5. The cluster that includes sequences sampled across several regions of Southeastern Asia (Hong Kong, South Korea, Taiwan, Singapore), as well as the major United Kingdom cluster (violet).

**Figure 1:**
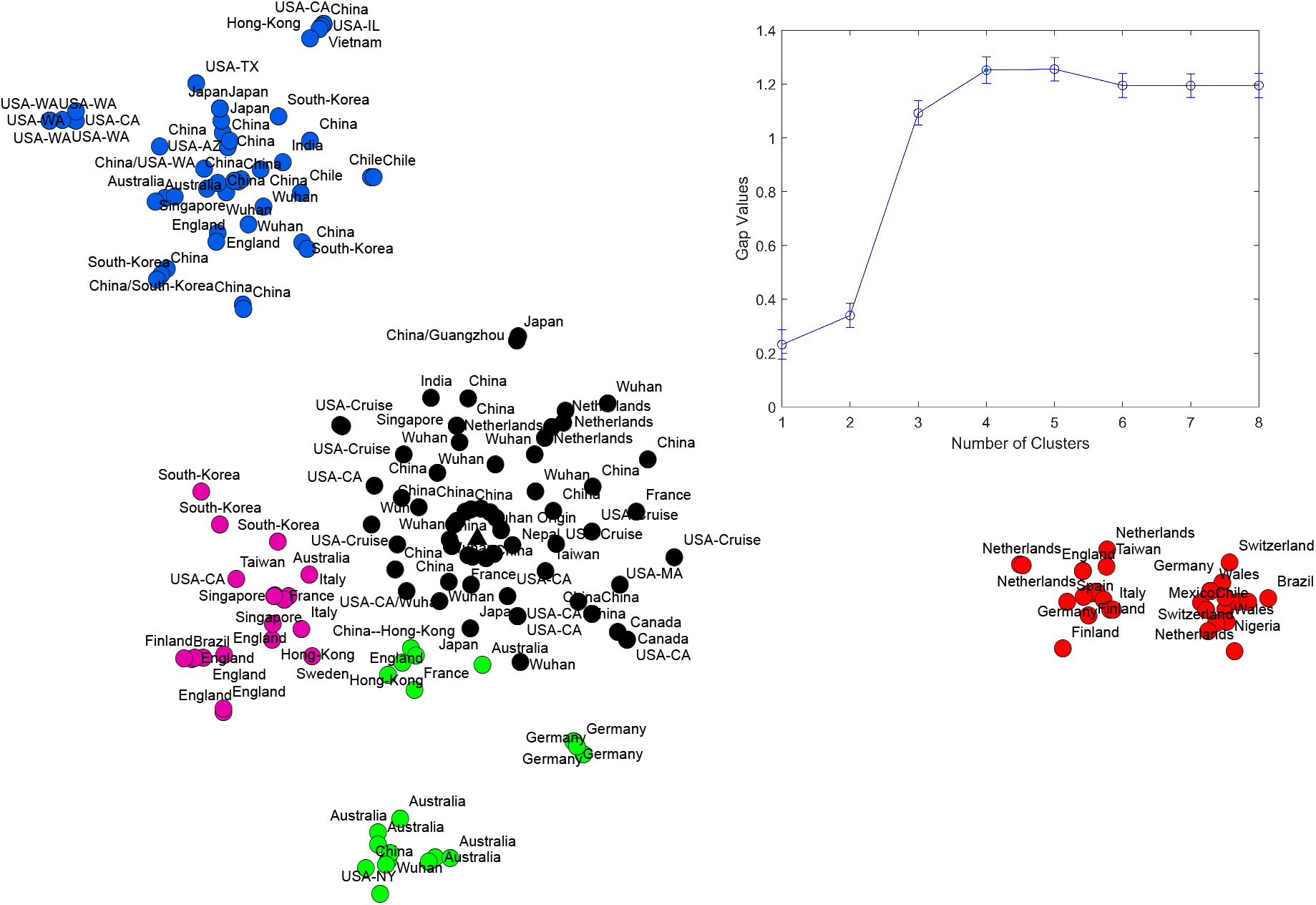
t-SNE plot of observed SARS-CoV-2 genomes. Wuhan-1 haplotype is depicted as a triangle. Identified clusters are highlighted in different colors. In the upper right corner, gap statistic values are shown.

It should be noted that these clusters do not consist exclusively of viral variants from the aforementioned regions, as air traffic probably drives the spread of SARS-CoV-2’s viral genomic variants from each cluster around the globe.

We observed the negative log-linear relationship between cluster sizes and maximum pairwise Hamming distances between the genomes inside the clusters (Fig. 3a, *R*^2^ = 0.984, *p <* 0.001). It suggest a preferential attachment-like mechanism of cluster formation, where the majority of newly appearing genomes tend to concentrate around the older genomes. This mechanism is further confirmed in the following analysis.

### 3.2 Transmission network

The transmission network produced by the algorithms described above is visualized in Fig. 2. Although the majority of viral haplotypes (80%) were sampled in a single geographical location, many of them were sampled in multiple locations. Such haplotypes are annotated on Fig. 2 by the full list of their sampling locations. In what follows, the most prevalent haplotype sampled in Wuhan and associated with the initial phase of the epidemic (highlighted in red on Fig. 2) will be referred to as Wuhan-1 haplotype. For the potential transmission links *e* discussed below, their bootstrapping-based probabilities *p*_*e*_ are reported.

**Figure 2:**
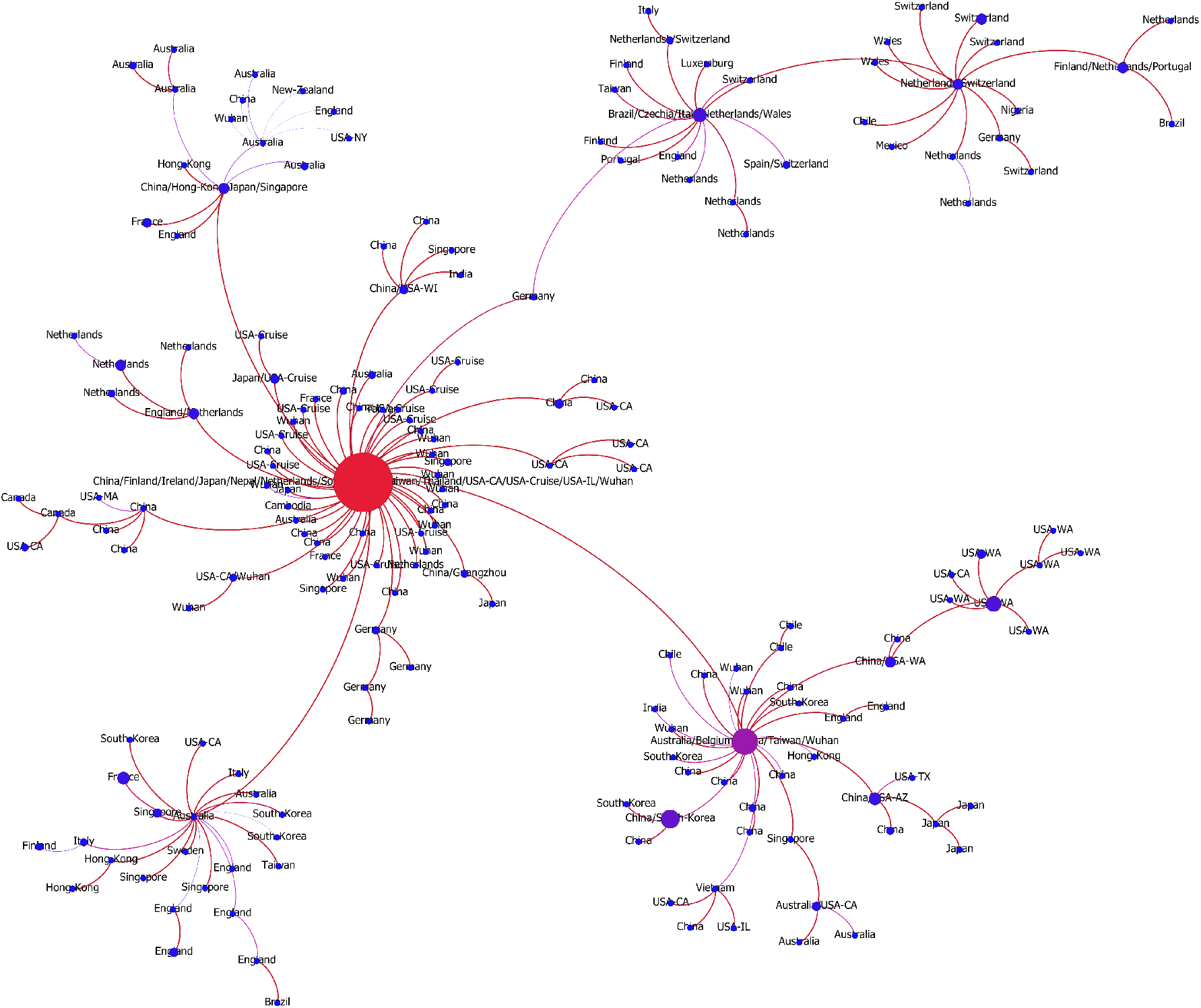
The transmission network constructed using SARS-CoV-2 genomes available as of March 10, 2020 and vizualized using Gephi [3]. Vertices represent viral genomes, and two vertices are connected by an arc if their mutational composition suggests potential direct or indirect transmission linkage between their hosts. Each vertex is annotated by the list of geographical locations were it was sampled. The thickness and color (from blue to red) of the edges are proportional to their bootstrapping probabilities.

#### 3.2.1 Network and temporal information

For the majority of genomes, their sampling times are known. The network structure was found to be in line with this temporal information, even though the network was constructed using the genomic data alone. Indeed, the correlation between network distances and differences in first sampling times between ancestor-descendant pairs of network nodes was 0.78 (*p <* 10^−116^). The fact that this correlation is not absolutely perfect is not surprising, as sampling times are much more prone to different sampling and reporting biases and may not accurately reflect actual transmission times. Even in such settings, for 86.10% of potential transmission pairs, their sampling times agree with each other, i.e. the potential source was sampled earlier than the potential recipient; and for 94.25% of these pairs their sampling times either agree or differ by up to 7 days. Finally, the mean minimum time difference between sampling times of potential transmission pairs was 3.74 days (95% *CI* = [2.75, 4.73]), while for the random pair of haplotypes the expected time difference was 20.48 days (95% *CI* = [20.21, 20.74]). This difference is statistically significant (*p <* 10^*−*45^, Kolmogorov-Smirnov test). The distribution of sampling time differences for linked pairs is shown in Fig. 3b.

**Figure 3:**
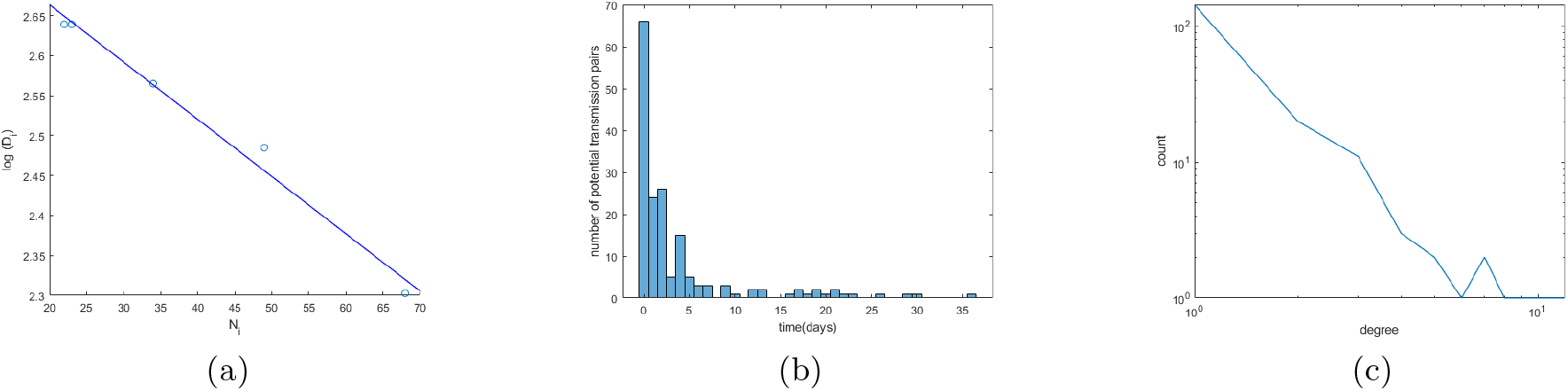
(a) Relationship between cluster sizes and the logarithms of maximum pairwise distances. (b) The distribution of sampling time differences for linked pairs. (c) Degree distribution of the transmission network (in log-log scale).

#### 3.2.2 Network structure

The transmission network is robust to an input data variation, with 97.34% of its edges being supported by the majority of bootstrap experiments, and 78.19% of edges having bootstrapping probabilities above 95%.

SARS-CoV-2 transmission network appears to be scale-free, with the the right-skewed degree distribution (Fig. 3c). Degree distributions of such networks follow power law (i.e. the probability of having a particular degree is proportional to the power of that degree), and they are often the result of a preferential attachment process, where a vertex joining a network gets connected to an existing vertex with the probability proportional to the degree of that vertex - the model is often described by the metaphor “the rich get richer”. Following [62, 29], we fitted the following distributions to the observed degree distribution of the transmission network: negative binomial, Yule, Pareto and Waring. To compare the goodness of fit yielded by different models, we used the Akaike information criterion (AIC) and Bayesian Information Criterion (BIC) (Table 1). The Pareto distribution, that represent the classical power-law demonstrated the best fit. The exponent of the Pareto distribution was estimated to be 1.20 (95%*CI* = [1.12, 1.34), indicating the higher tendency of vertices to be connected to hubs (high-degree vertices).

**Table 1:** Comaprison of different models for the transmission network degree distribution.

| Distribution | AIC | BIC |
| --- | --- | --- |
| Negative Binomial | 708.49 | 714.96 |
| Pareto | 603.48 | 609.95 |
| Waring | 704.59 | 711.06 |
| Yule | 804.91 | 808.15 |

Furthermore, the correlation between vertex degrees and sampling frequencies of the corresponding genomes was high: *ρ* = 0.8932, *p <* 10^*−*65^. All these observations suggest that few genomes were responsible for the majority of possible transmissions.

#### 3.2.3 Transmission history

In general, the structure of the potential transmission network agrees with the distribution of t-SNE clusters (Fig. 1) and allows to hypothesize multiple transmission routes. In particular, the virus spread is characterized by multiple introductions of SARS-CoV2 into regions and countries: at this point for 14 out of 34 countries with reported sequences multiple introductions could be claimed, although additional data could adjust these estimations. Below we summarize the information about the transmission pathways in different regions outside the mainland China (whose subnetwork is depicted on Fig. A1) that could be deduced from the inferred network. It should be noted that we are currently lacking whole-genome sequencing data from Iran and Eastern Europe, therefore we concentrate on the analysis of regions with a sufficiently large number of available samples.

**USA** (Fig. A2) At this point, there are indications of multiple introductions of SARS-CoV2 into the country (not counting the cases from the Grand Princess cruise ship), as well as of sustained human-to-human transmissions inside the country. Most of introduced haplotypes could be directly linked to the first epidemic wave in mainland China, with the average graph distance between US haplotypes (Washington cases excluded) and Wuhan-1 haplotype being equal to *d* = 1.79. In particular, the state of California alone could have exhibited multiple introductions with no identified significant clustering of cases, as its observed viral haplotypes were either also sampled in China (2 cases) or linked directly to the haplotypes in mainland China (2 cases, *p*_*e*_ = 1), or to haplotypes from Singapore, Vietnam, Australia and Canada (4 cases, *p*_*e*_ = 0.98, 0.97, 0.99 and 1, respectively) which are, in turn, linked to haplotypes sampled in mainland China (*p*_*e*_ = 0.98, 0.77, 1 and 1). In contrast, the haplotypes from the state of Washington form a connected subtree and thus suggest a single introduction from mainland China (since the root of the Washington subtree was also sampled in China) followed by the sustainable human-to-human transmissions inside the state (mean *p*_*e*_ = 0.99). In addition, the network suggests independent introductions to Massachusetts, Wisconsin, New York, Illinois and Arizona. So far, two possible cases of virus transmission between US states could be identified: from Arizona to Texas (*p*_*e*_ = 0.83) and from Washington to California (*p*_*e*_ = 1). Finally, as expected, the sequences from the Grand Princess cruise ship could be linked to a single case identical to the Wuhan-1 haplotype.

##### Western Europe

The major European cluster is linked to the Wuhan-1 haplotype through the haplotype sampled in Germany on January, 28, 2020 (*p*_*e*_ = 0.79) (Fi. A3). The parent of this haplotype is the Wuhan-1 haplotype (*p*_*e*_ = 0.91) and its only child is the haplo-type later sampled in Italy. This potential transmission route is in agreement with epidemiological and molecular evidence reported by other sources [4, 22, 23]. For Italy, the analysis suggests that there was another independent SARS-CoV-2 introduction with no genomic evidence that it led to further spread. Similarly, at least two introductions are hypothesized for Netherlands (Fig. A4); however in that case both resulted in sustained host-to-host transmissions inside the country. The first Netherlands cluster is part of the major European cluster, while the second one could be linked to the Wuhan-1 haplotype (*p*_*e*_ = 1). Interestingly, this cluster has the genetic signature in the form of codon deletion in nsp2 genomic region that was observed only there; furthermore, both viral substrains are not geographically separated and co-exist in the same cities. A similar situation is observed in Germany, connected to the major European cluster and a separate branch sampled at North Rhine-Westphalia and directly linked to the Wuhan-1 haplotype (*p*_*e*_ = 1). Multiple introductions have also been observed in Finland. Haplotypes from Switzerland, Spain and Czech Republic are only observed in the main European cluster and most probably were introduced from Italy; in the latter case, this claim has an epidemiological support as the infected person reportedly had traveled Italy.

The epidemiological history in the United Kingdom seems to be quite different from that of continental Europe (Fig. A5). There are multiple separate clusters detected there, three of which cannot be associated with other European cases, but are directly linked to the sequences sampled in China and Australia in January, 2020 (mean *p*_*e*_ = 0.82). Regarding the remaining haplotypes, one belongs to the second Netherlands cluster, while the remaining belong to the major European cluster (the majority of them being sampled in Wales). Two clusters show indications of intra-country transmissions (mean *p*_*e*_ = 0.93).

##### East Asia

All introductions in Singapore, Japan and South Korea (Figs. A6, A7 and A8) were linked to the haplotypes observed in mainland China. Singapore possibly experienced four such introductions (mean *p*_*e*_ = 0.99). In both Japan and South Korea, two potential introductions could be predicted, one linked to Wuhan-1 haplotype, and the other linked to intra-country transmissions (mean *p*_*e*_ = 0.99 in Japan and *p*_*e*_ = 0.86 for South Korea).

##### Australia

There are indications of at least three potential virus introductions to Australia either from mainland China or, as discussed in the previous subsection, from Iran, although the latter claim is currently based only on epidemiological evidence. Three of the corresponding clusters have evidences of intra-country transmissions (mean *p*_*e*_ = 0.88).

##### Central and South America and Africa

These regions currently reported few SARS-CoV-2 haplotypes, and the majority of cases most probably represent the third wave of the epidemics. Indeed, viral variants sampled in Nigeria, Mexico, 2 variants from Brazil and 1 variant from Chile are linked to haplotypes from the major European cluster (mean *p*_*e*_ = 1) and have reported travel history to Italy, while one variant from Brazil is linked to the genome from the United Kingdom (*p*_*e*_ = 0.99), thus supporting the hypothesis that the virus was imported from continental Europe. On the other hand, three other Chilean haplotypes belong to the Pacific cluster and could be linked to the haplotype sampled in mainland China, Taiwan, Australia and Belgium (mean *p*_*e*_ = 0.92).

## 4 Discussion

In this work, we report the results of molecular surveillance analyses of SARS-CoV-2 prior to the transition from epidemic to pandemic state. Our aim was to identify and analyze the transmission pathways that allowed the epidemic to rapidly progress from the initial outbreak in Wuhan to the pandemic that is now affecting almost every geographic region of the globe [14, 32, 33]. To achieve this aim, we proposed, implemented and used a computational framework to recover the network of potential SARS-CoV-2 transmissions using the aggregated genomic data retrieved from GISAID repository [56]. The analysis allowed to identify the potential transmission links and routes of disease introduction in different regions of the planet, and confirm the hypothesis that there were multiple sources of introduction to the majority of countries. This conclusion supports the implementation of travel restrictions and border screening which have already been implemented in different locations. It should be also emphasized that the fact that a number of countries exhibited multiple introductions demonstrates how different susceptible human subpopulations are interconnected nowadays. This allowed the virus to exploit multiple transmission pathways and spread so rapidly. At the same time, our work underscores the need to put in place coordinated efforts involving multiple countries in order to achieve epidemic control. The scale-free structure of the global SARS-CoV-2 transmission network suggests that a few genomes were responsible for the spread of the virus. The question whether this pattern is due to founder effects, epidemiological settings or differences in phenotypic features across SARS-CoV-2 genomic variants remains open and will require further investigation. Furthermore, scale-freeness of the network should be taken into account in epidemiological modelling and planning of public health intervention measure, since epidemic processes on such networks exhibit a specific behaviour [44, 50].

The ongoing pandemic of SARS-CoV-2 is the first global public health emergency for which next-generation sequencing technologies have been employed at such scale. This has led to a high density sampling that is unprecedented for both the geographic extent of virus spread and the evolutionary space explored by the virus since its emergence in China. Since the pandemic is currently only a few months old, no epidemiologically relevant lineages from the beginning of the epidemics are expected to die out anytime soon. Indeed, although the virus genetic diversity is gradually increasing, particular genomic variants continue to be repeatedly sequenced at different time points and geographical locations. It provides the means of tracking virus spread and evolution across time and space from the beginning of the epidemics using the methods of computational genomics and molecular epidemiology, and make conclusions about the potential routes of transmission.

We should warn that the results of such molecular surveillance analyses should be interpreted with caution. First of all, the genomic analyses do not necessarily replace traditional epidemiology methods that aim to investigate sources of transmission based on traditional surveillance systems that keeps track of the trajectory of the epidemic. Rather, genomic analyses complement and confirm other epidemiological findings using the sequencing data as an independent source of information that is not subject to the biases associated with the traditional epidemiological data [27, 2]. Second, it is important to understand that the edges in the estimated global transmission network may not be synonymous with actual transmission events, but rather link infected hosts from the same epidemiological transmission clusters. Furthermore, the analysis is subject to the limitations associated with the nature of genomic data that include a small amount of cases in the beginning of the epidemics, underreporting and potential multiple sources of epidemic introductions. Further, the dataset available for this analysis is a convenience sample rather than a random sample within infected individuals, which results from the aggregation of data from different countries and sequencing labs and instruments. This is an inevitable consequence of sequencing data analysis since the procedure itself can be relatively expensive when implemented on a large scale [57, 55, 45], and the decision to sequence each particular case is largely done subjectively in each specific country and lab.

In response to the rapidly growing number of cases of COVID-19 the authorities in different countries around the world have implemented unprecedentedly stringent travel and movement restrictions. Those measures were taken separately and independently country by country with different levels of escalation and at different times. In this context it is important to emphasize the importance of globally coordinated measures and collaborations that should be supported by timely and evidence-supported analysis of reliable epidemiological data of diverse nature. Automatic high-performance computing-based molecular near real-time surveillance systems such as Nextstrain [28], HIV-Trace [36] and GHOST [40] could be instrumental in such public health global surveillance and decision making.

## Data Availability

Genomics data and associated metadata for this study have been obtained from the Global Initiative on Sharing All Influenza Data (GISAID) database. The scripts for network reconstruction are available at https://github.com/compbel/SARS-CoV-2.

## Acknowledgements

We acknowledge all reserachers and laboratories who contributed their data to GISAID database. A full listing of all originating laboratories and authors is available at https://www.dropbox.com/s/4wjpi9lv5tlnsxh/gisaid.xlsx?dl=0

## Software and data availability

The scripts for network reconstruction are available at https://github.com/compbel/SARS-CoV-2. The results of our analysis are posted at https://publichealth.gsu.edu/research/genomic-coronavirus/

## Authors’ Contributions

PS designed and implemented algorithms, processed and analyzed the data and wrote the paper; AK analyzed the data and wrote the paper; PIB designed algorithms and processed the data; AZ designed the algorithms and wrote the paper; GC analyzed the data and wrote the paper. All authors read and approved the final manuscript.

## Appendix

Below are transmission subnetworks for the regions described in the paper.

**Figure A1:**
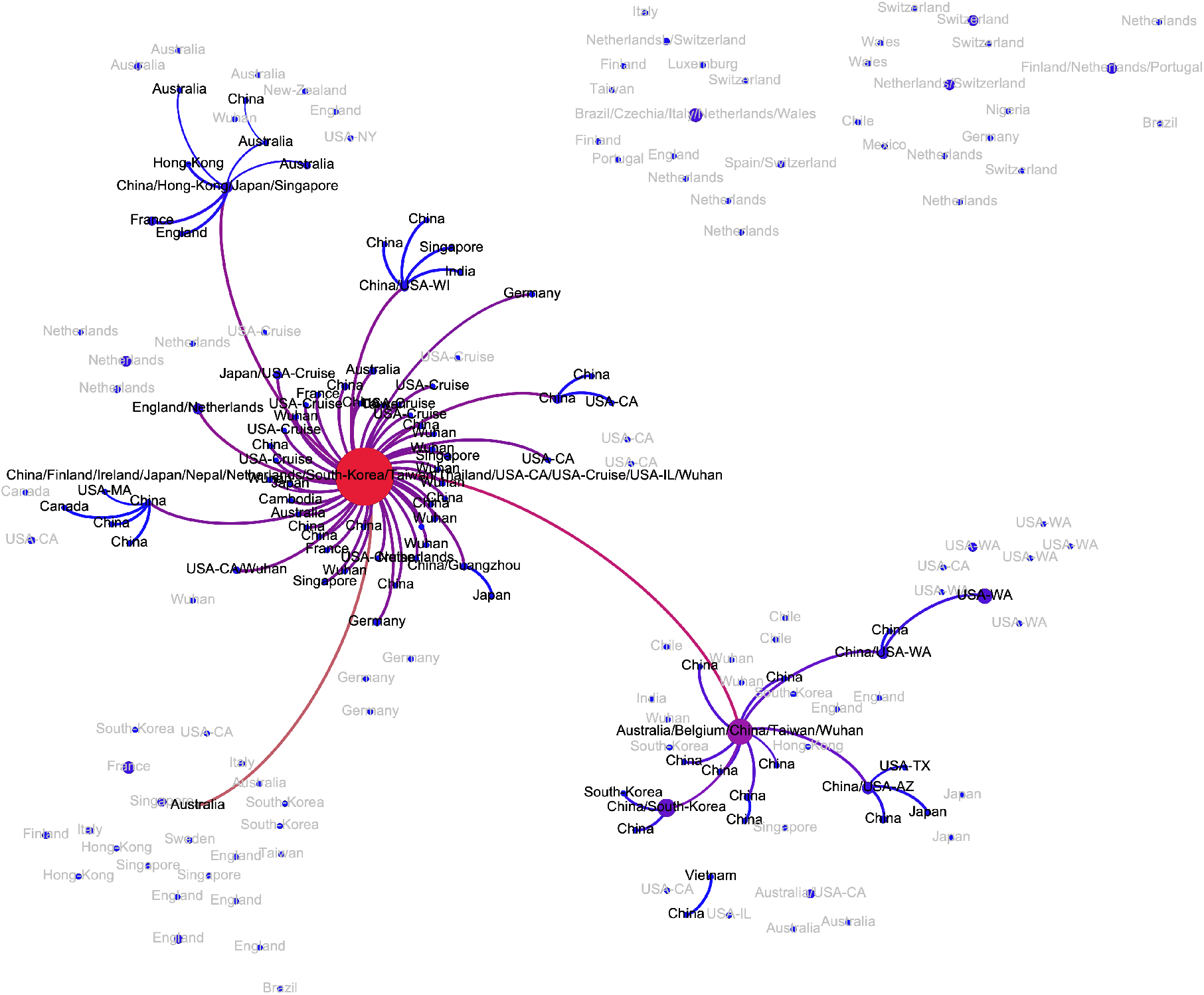
Transmission subnetwork of genomes sampled in China.

**Figure A2:**
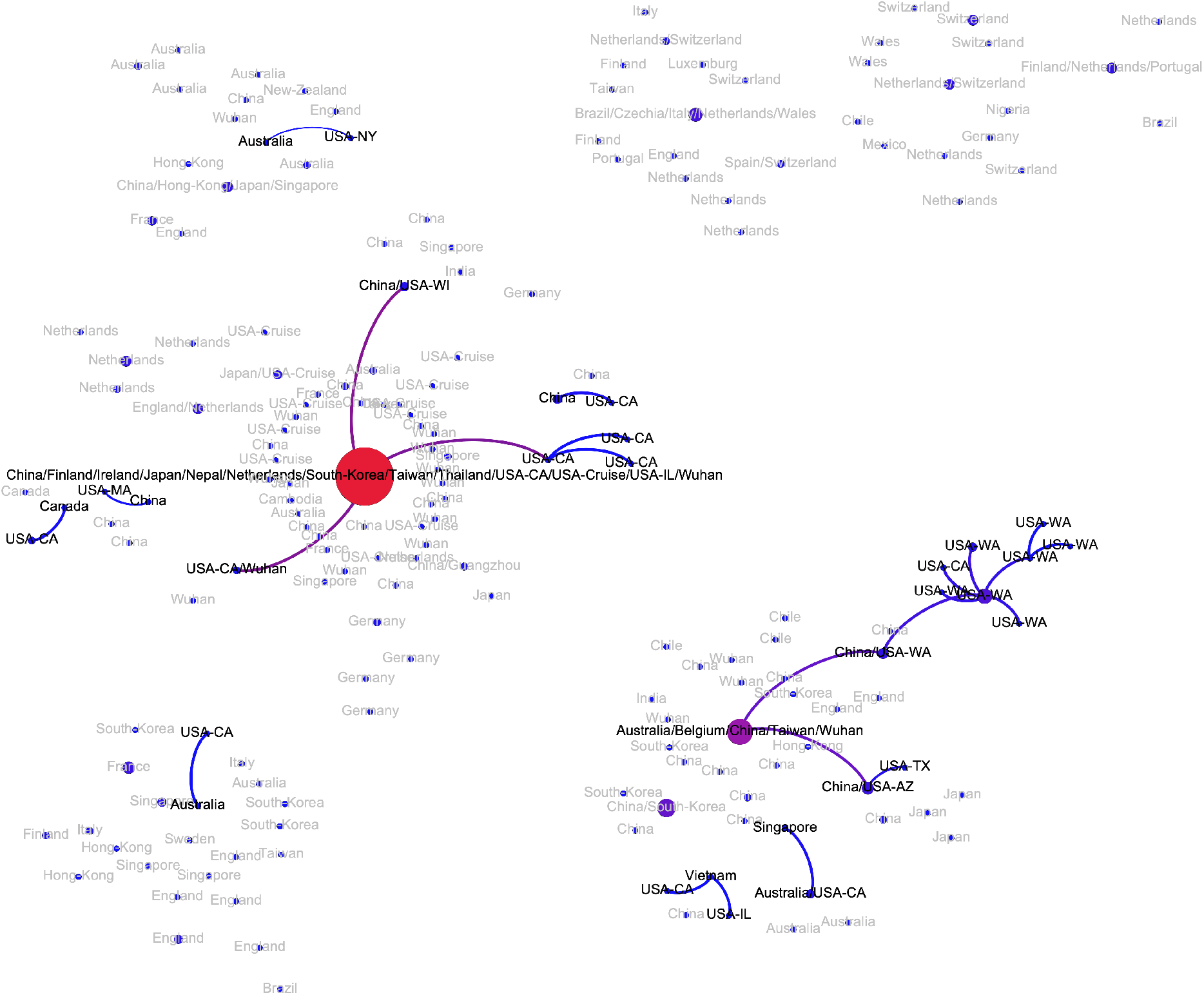
Transmission subnetwork of genomes sampled in USA.

**Figure A3:**
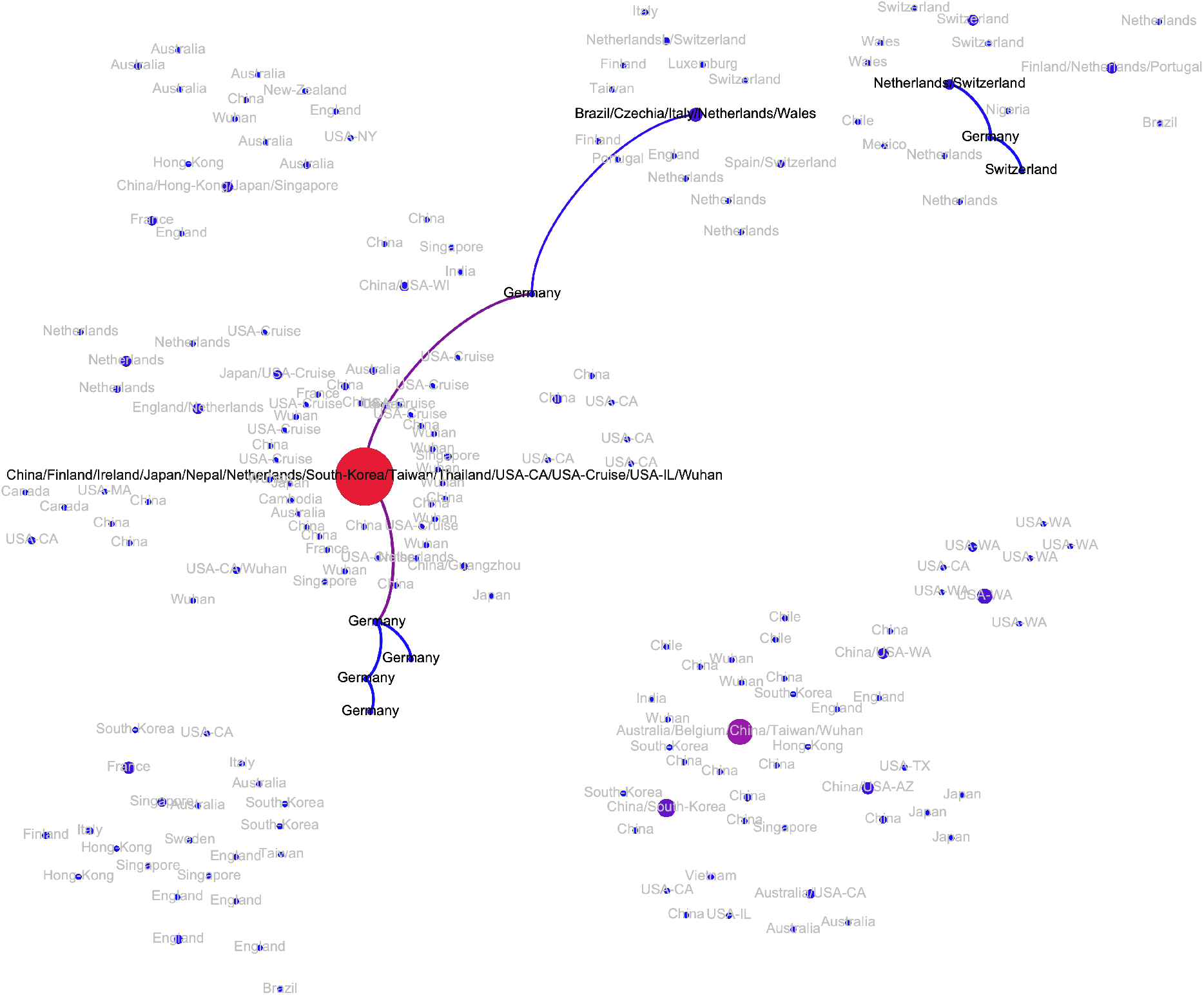
Transmission subnetwork of genomes sampled in Germany.

**Figure A4:**
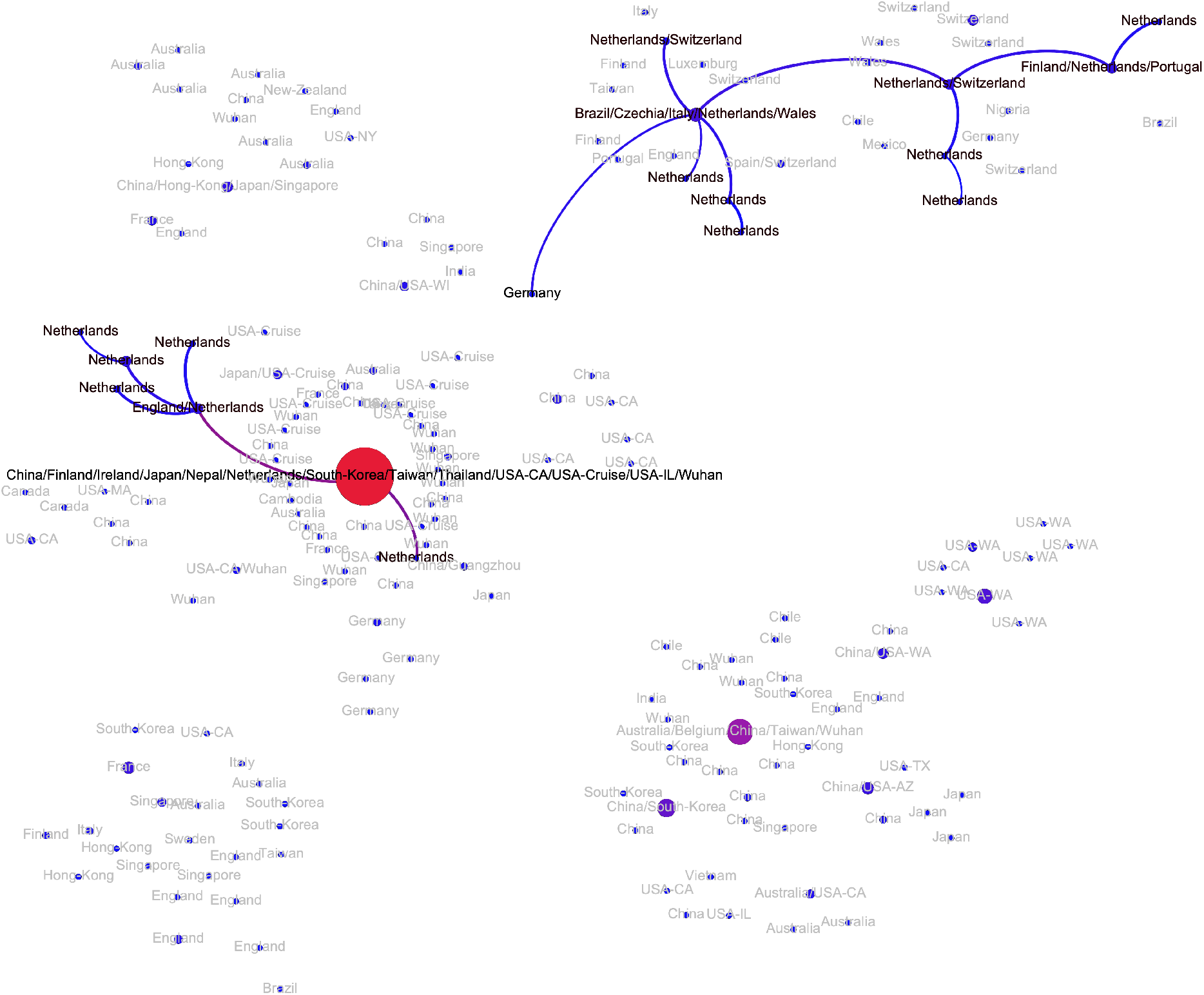
Transmission subnetwork of genomes sampled in Netherlands.

**Figure A5:**
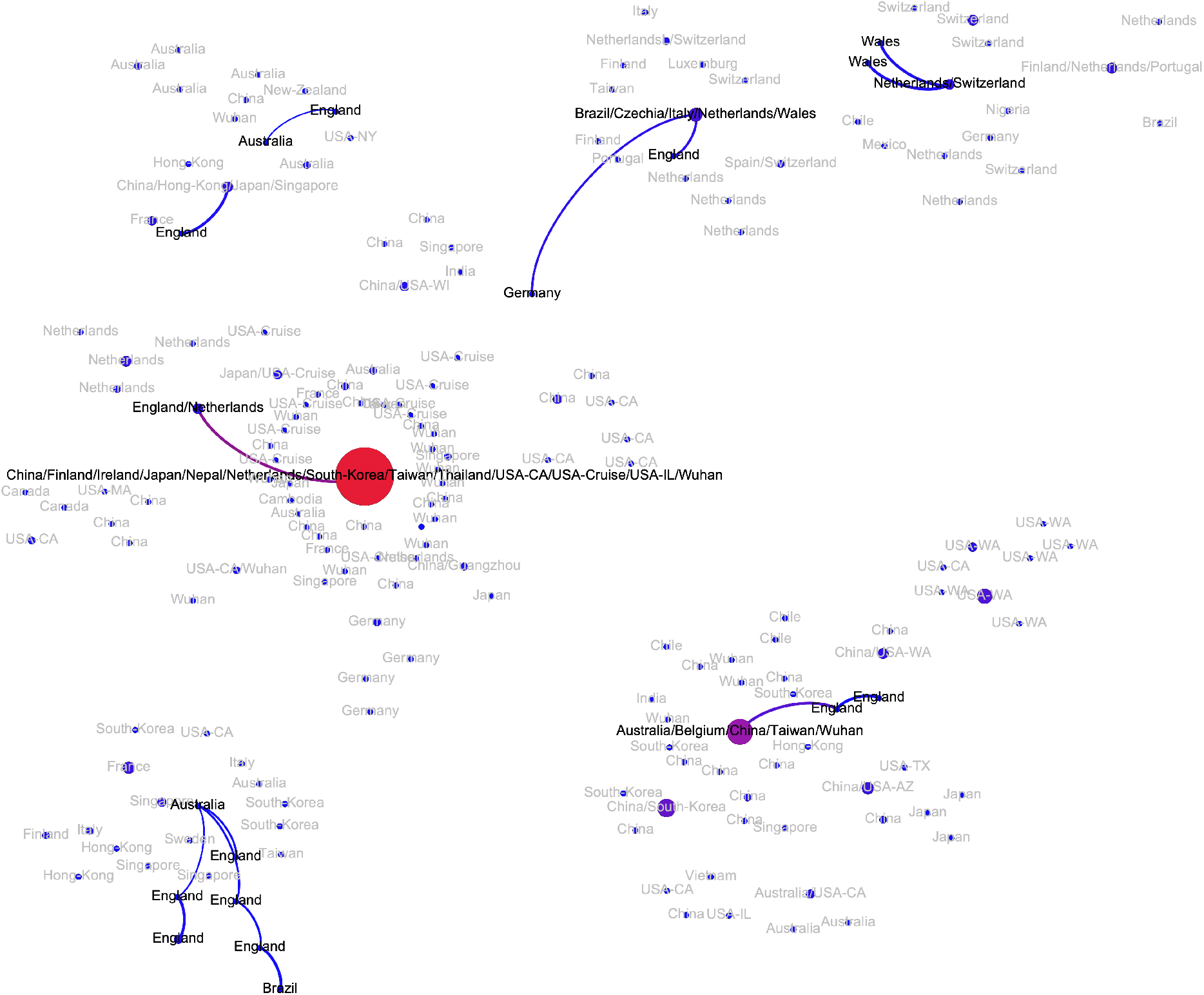
Transmission subnetwork of genomes sampled in UK.

**Figure A6:**
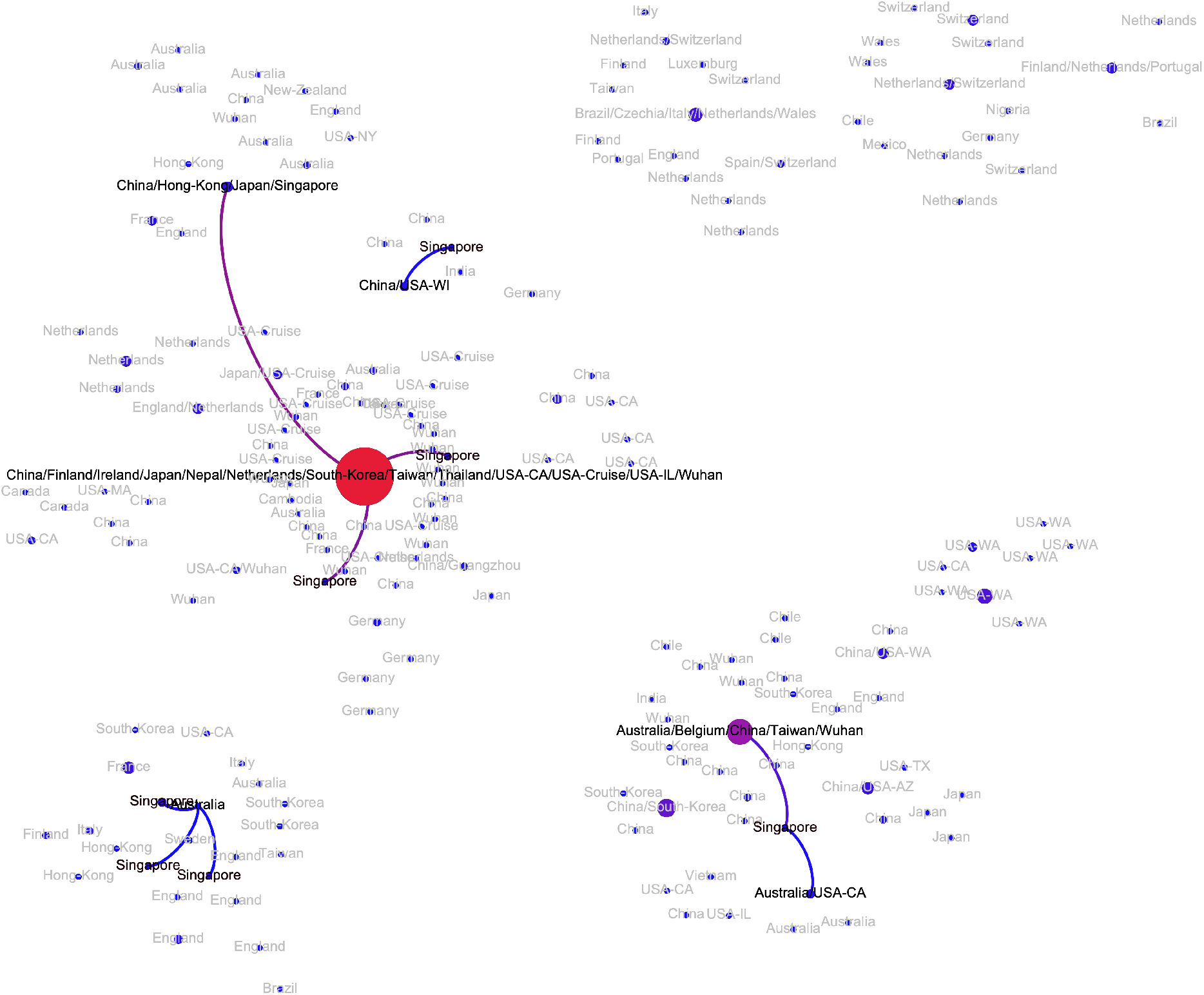
Transmission subnetwork of genomes sampled in Singapore.

**Figure A7:**
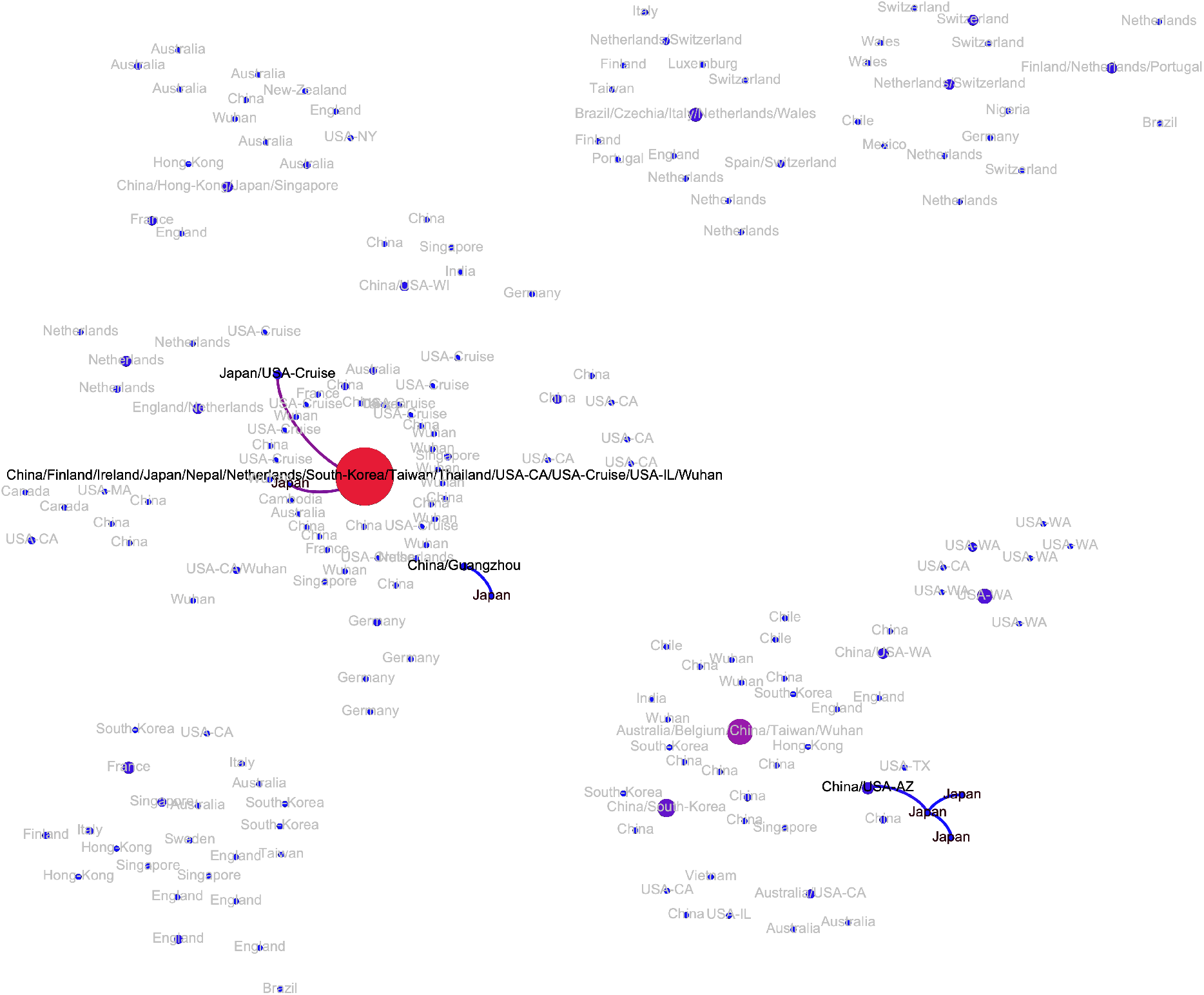
Transmission subnetwork of genomes sampled in Japan.

**Figure A8:**
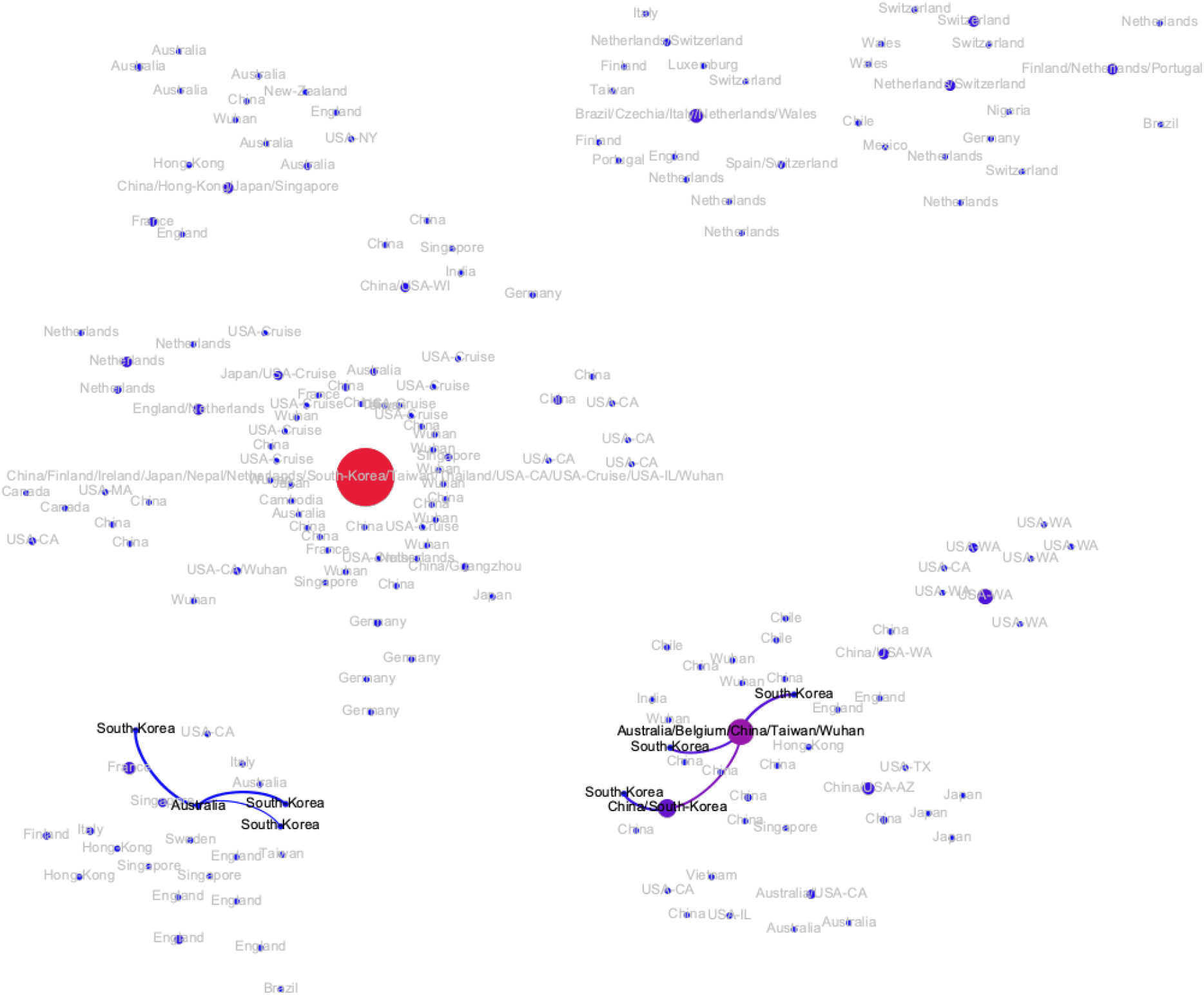
Transmission subnetwork of genomes sampled in South Korea.

**Figure A9:**
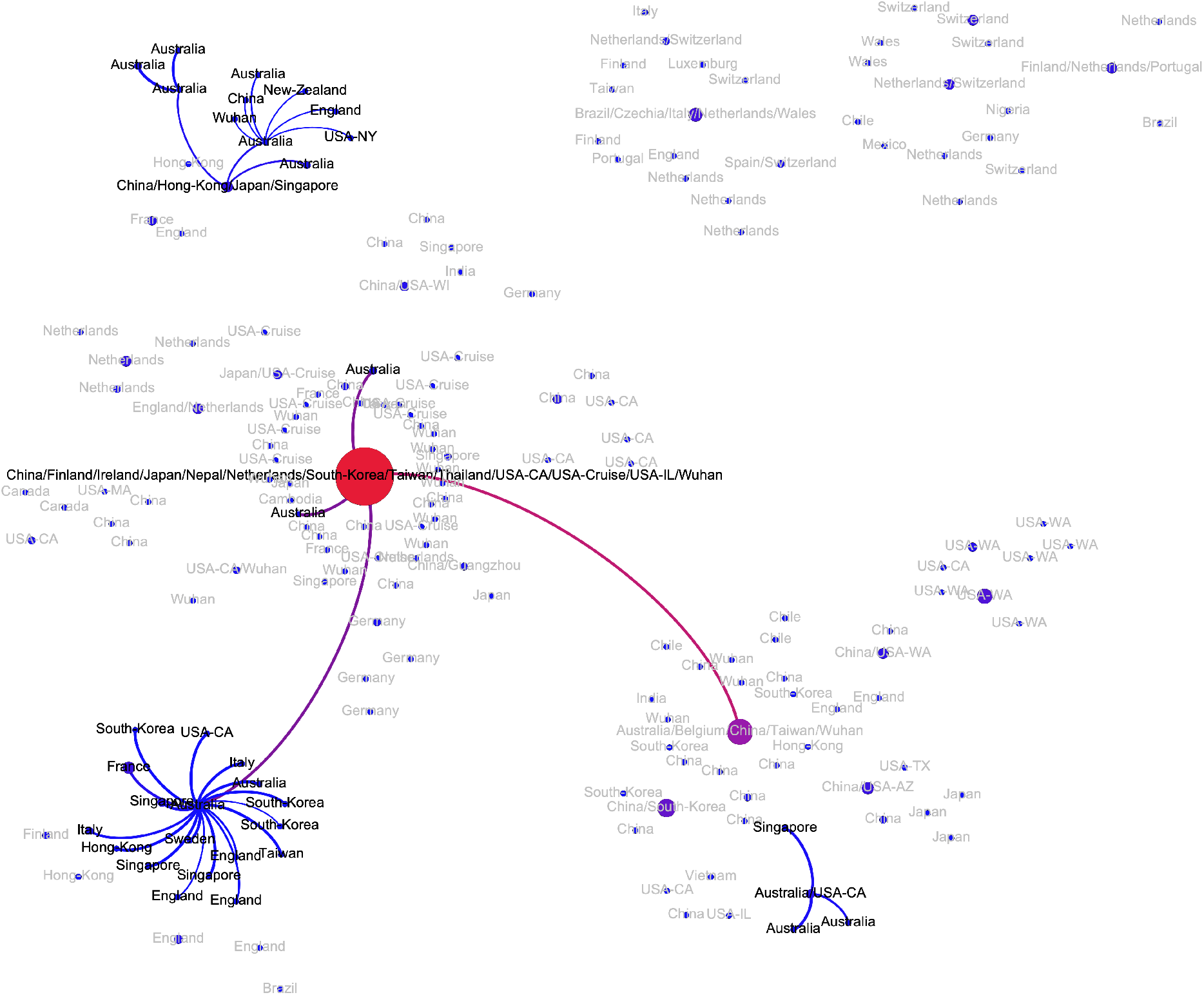
Transmission subnetwork of genomes sampled in Australia.

**Figure A10:**
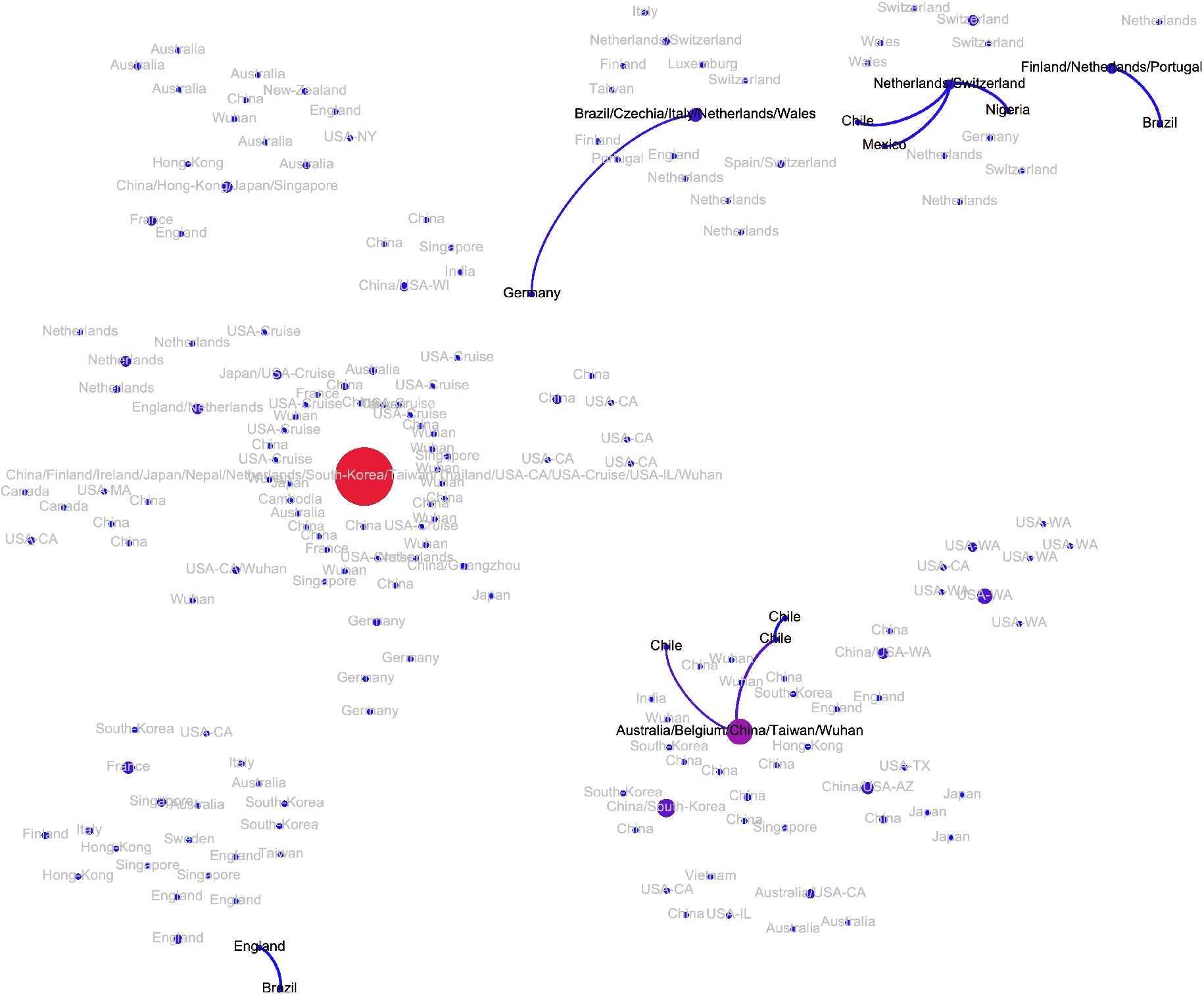
Transmission subnetwork of genomes sampled in Africa, Central and South America.

